# Myo-Inositol Concentration in the Medial Prefrontal Cortex is Associated with Changes in Brain White Matter Microstructure in Early Psychosis

**DOI:** 10.1101/2025.08.15.25333577

**Authors:** Tommaso Pavan, Qiaochu Wang, Yasser Alemán-Gómez, Raoul Jenni, Martine Cleusix, Luis Alameda, Kim Q. Do, Philippe Conus, Patric Hagmann, Pascal Steullet, Paul Klauser, Lijing Xin, Ileana Jelescu

**Author notes:** equal contribution (Lijing Xin, Ileana Jelescu). Corresponding author: Tommaso Pavan; address: Centre de Recherche en Radiologie, PET3, CHUV, Rue du Bugnon 46, 1011, Lausanne, Switzerland.

## Abstract

Recent research highlights the critical role of white matter (WM) alterations in psychosis and schizophrenia (SZ), reporting volumetric and structural brain changes in affected individuals. In this study, we explored the role of astroglia in SZ, which is believed to play a role in white matter integrity. We investigated for the first time the associations between advanced diffusion Magnetic Resonance Imaging (dMRI) measures of WM microstructure, and Magnetic Resonance Spectroscopy (MRS)-derived glial markers in 30 subjects with early psychosis (EP, 24±6 y.o.) versus 41 healthy controls (HC, 25±6 y.o.).

We focused on two metabolites involved in glia: myo-Inositol (myo-Ins) and total Choline (tCho), measured in the medial prefrontal cortex (mPFC), relating them to quantitative dMRI metrics derived from Diffusion Kurtosis Imaging (DKI) and WM Tract Integrity-Watson (WMTI-W) biophysical model including mean diffusivity and kurtosis, axonal water fraction and extra-axonal diffusivities in the whole white matter.

Our findings reveal a difference between EP and HC in WM diffusivities, specifically in extra-axonal parallel direction, but not in MRS metabolites. However, we found that the mPFC myo-Ins concentrations in EP are exclusively and strongly associated with proximal WM microstructure features, in the form of a positive correlation with axonal water fraction, a proxy for axonal density, and negative correlation with extra-axonal parallel diffusivity, suggesting the white matter alteration could be linked to astrocytic changes in subset of early psychosis.

## Introduction

Psychotic disorders, including schizophrenia (SZ), pose considerable challenges for the affected individuals, their families, and the broader community^1^. Recent research highlights the key role of white matter (WM) in this disease, reporting global structural changes across the brain of people suffering from psychosis^2,3^. Thanks to its ability to exploit the random motion of water molecules, diffusion MRI (dMRI) allows us to explore the cellular environment and infer the microstructural properties of the underlying biological tissue in vivo^4^. In participants with psychosis, Diffusion Tensor Imaging (DTI) studies of WM consistently reported patterns of reduced fractional anisotropy (FA) and increased mean diffusivity (MD)^3,5,6^. These WM alterations are considered global, affecting the entire WM to varying degrees depending on the region^7^, most notably in the frontal and prefrontal areas^2,3^. One of the largest meta-analysis of schizophrenia patients vs controls^3^ reported the highest effect size in dMRI metrics for average FA across the whole brain (*Cohen’s d*=0.42), followed by FA in the anterior corona radiata, and the genu and body of the corpus callosum. In our recent work^8^, we reproduced these high effect sizes in the same brain regions, in patients with early psychosis (EP) and SZ. Additionally, we reported a decrease in Diffusion Kurtosis Imaging metrics^9^ (DKI, an extension of DTI which provides complementary information about tissue heterogeneity) in both clinical groups as compared to age range-matched controls, and identified specific microstructure patterns of alterations using the biophysical diffusion model, White Matter Tract Integrity - Watson^10^ (WMTI-W). Specifically, WMTI-W enables the estimation of compartment-specific (intra- and extra-axonal) properties that are excellent proxies for intra-axonal injury, inflammation and abnormal myelin integrity^11–13^.

Thanks to the advanced dMRI techniques, we found that WM alterations manifested predominantly in the extra-axonal space, as a significant increase in extra-axonal diffusivities, and that they were already present at the EP stage^8^. This latter finding is consistent with literature showing that WM changes have been observed before the onset of psychosis^2,14^. In ultra high-risk individuals, existing WM alterations do not appear to suddenly worsen with the onset of psychosis^14^. Instead, studies show a progressive reduction in WM FA in those who transitioned^15^, and a prematurely arrested FA increase during development in a longitudinal context^16^. Moreover, these alterations were not associated with symptomatology^3,8^.

Various mechanisms have been proposed to explain WM alterations, ranging from early aberrant pruning^17^ and neuroplasticity^18^, to redox dysregulation^19^, and glutamatergic disfunctions^20,21^. A growing number of studies included neuroinflammation^22–24^ and changes in neuroglia^17,25^ in the etiology of psychosis. *Post-mortem* studies reported a decrease in astrocytes^26^ and oligodendrocytes^27^, which were estimated to be 20 to 27% lower than in controls^28,29^, in SZ specimens when compared to controls. On the other hand, in those specimens who had high expression of inflammatory markers, ~40-50% of the brains exhibit increased immune activation (as measured by interleukin-6) in the dorsolateral prefrontal (PFC) and orbitofrontal cortices^30^, increased astrogliosis^31^ and reduced gray matter volume^32^. In-vivo dMRI studies have reported links between WM alterations (FW, free-water imaging^33^, i.e. reduced tissue compartment) and increased blood inflammatory cytokines^34^.

Studies using Magnetic Resonance Spectroscopy (MRS), a quantitative MR technique that measures the brain tissue metabolic composition in vivo, have primarily focused on Myo-Inositol (myo-Ins) and total Choline (tCho) levels in patients^35^ to investigate glial changes in SZ. Myo-Ins plays three main roles, as constituent of the lipids (phosphoglycerides) in biomembranes, as regulator of the cellular volume through osmolytic action, and as part of intracellular second messenger system^36^. Reports indicate elevated expression of myo-Ins in astrogliosis^37^, glioma^38^ (independent of membrane turnover), and the myo-Ins diffusivity seems associated to astrocyte hypertrophy in mice^39^. Thus, myo-Ins has been proposed as a marker of astrocyte density^38^ or integrity^40^, and it was associated with inflammatory processes when increased.

In SZ, medial prefrontal myo-Ins is often found to be reduced with a small effect size (standardized mean difference = 0.19)^41^, and these reductions are evident already in the early, untreated stages of psychosis^40^. Decreased myo-Ins has been associated with higher depressive symptoms in SZ^42^ and across various psychiatric diseases^43^, while an increase in myo-Ins to levels closer to HC improves general symptomatology and social-occupational functioning in first-episode SZ^40^. On the other hand, higher myo-Ins levels have been reported in treatment-naїve first-episode psychosis compared to HC^44^, in treatment resistant compared to both non-treatment resistant SZ and HC, and associated with positive symptoms^44^. These findings may suggest that antipsychotic treatment may decrease the myo-Ins levels^45^, which seems to be higher only during an active/untreated phase.

Choline is utilized for phospholipid synthesis and strongly correlates with cell density, making it a marker for membrane turnover^36,46^ and myelin^36^. Similarly to myo-Ins, tCho concentration is higher in (astro and micro) glia than neurons^47^, in oligodendrocytes precursor cells^48,49^, and its diffusion has been proposed as marker of (micro) glia morphology^49^. tCho is reported to increase in dorsolateral and medial PFC in SZ compared to HC^50,51^, in treatment-naїve first-episode psychosis^44^, and in treatment resistant SZ compared to non-treatment resistant SZ and HC^45^, likewise myo-Ins.

In this study, we aim to investigate the role of astroglia underlying WM alterations in psychosis by linking concentrations of metabolites associated with glial changes (myo-Ins and tCho) in the medial prefrontal cortex (mPFC), a key area for SZ^3^, to the WM microstructure changes observed in early psychosis. To our knowledge, only one other study^42^ investigated a similar association in chronic SZ, and reported that, when correlating metabolites with dMRI measures of the anterior corona radiata, myo-Ins was negatively associated with FA in both EP and HC, whereas tCho was not, and that myo-Ins was the sole metabolite associated to WM diffusion changes when accounting for other major metabolites (NAA, Glu).

Based on these findings and on previous literature, we anticipate finding lower myo-Ins levels in our treated EP cohort as compared to HC. Therefore, we expect a negative association of myo-Ins with DTI diffusivities and extra-axonal diffusivities, and a positive association with kurtosis and axonal water fraction (i.e. lower glial density leads to higher extra-axonal water mobility, lower tissue heterogeneity and lower density of axons and other cellular processes).

In addition, to test the specificity of this association to myo-Ins and tCho, we examine the association with three other major metabolites in an exploratory and comparative manner: glutamate (Glu) and glutathione (GSH), which are respectively involved in glutamatergic dysfunction^20,21^ and oxidative stress^19^, and N-acetylaspartate (NAA), a marker for neural metabolism and myelin formation^52,53^, that has been associated with WM alterations. Furthermore, we hypothesize that WM microstructure metrics will correlate with the glial marker rather than neuronal metabolites since most of our previous findings indicate a marked increase in extra-axonal diffusivities^8^, consistent with glial density changes that would reduce water hindrance outside the axons.

In previous work^8^, we also tested the hypothesis that oxidative stress^19^ estimated from blood markers could be partially responsible for the WM alteration in SZ. Here, we briefly tested again this hypothesis using direct brain MRS GSH.

## Methods

### Participants

Data were collected from 79 individuals (**Table 1**) divided into two groups: 49 healthy controls (HC) and 30 participants with a diagnosis of EP, i.e. individuals within 5 years of having met psychosis threshold according to the Comprehensive Assessment of At-Risk Mental States^57^, CAARMS. Subjects with psychosis related to intoxication or organic brain disease, IQ*<*70, reporting alcoholism, drug abuse, major somatic disease, documented anamnestic or current organic brain damage were excluded. EP subjects were recruited from the Treatment and Early Intervention in Psychosis Program^58^ (TIPP, University Hospital of Lausanne, Lausanne, Switzerland). HC participants were recruited from the same sociodemographic area as the patients. HC participants were excluded if they or a first-degree family member reported to have suffered from neurological, traumatic, major mood disorder, psychosis, prodromal symptoms, and current or past antipsychotic treatments. The study was approved by the local Ethics Committee (CER-VD 382/11 and 2018-01731).

**Table 1.** Cohort demographics. EP: early psychosis; HC: healthy controls; mPFC: medial prefrontal cortex; GSH: glutathione; Glu: Glutamate; Ins: Inositol; TCho: Total Choline; WM: white matter; GAF: global assessment of functioning; PANSS: positive and negative syndrome scale; CPZ: chlorpromazine. ^†^: brain volumes t-tests statistics were computed on the volumes normalized by the estimated total intracranial volume (eTICV).

|  |  | EP (N=30) |  | HC (N=49) |  | EP-HC |  |
| --- | --- | --- | --- | --- | --- | --- | --- |
|  |  | Mean | Std. Dev. | Mean | Std. Dev. | t-value | p-value |
| Age (years) |  | 24.58 | 5.27 | 25.58 | 4.93 | -0.85 | - |
| myo-Ins<sub>mPFC</sub> (μmol/g) |  | 7.91 | 1.13 | 8.23 | 0.94 | -1.31 | - |
| tCho<sub>mPFC</sub> (μmol/g) |  | 2.56 | 0.44 | 2.54 | 0.33 | 0.13 | - |
| Glu<sub>mPFC</sub> (μmol/g) |  | 13.49 | 1.40 | 13.91 | 1.23 | -1.37 | - |
| GSH<sub>mPFC</sub> (μmol/g) |  | 1.25 | 0.30 | 1.25 | 0.27 | 0.07 | - |
| NAA<sub>mPFC</sub> (μmol/g) |  | 12.59 | 1.34 | 12.86 | 0.99 | -0.96 | - |
| Brain volume (cm<sup>3</sup>) |  | 1162.22 | 97.74 | 1177.89 | 115.43 | -2.8 | 0.0068<sup>†</sup> |
| GM volume (cm<sup>3</sup>) |  | 680.81 | 60.43 | 688.05 | 66.70 | -1.53 | - |
| WM volume (cm<sup>3</sup>) |  | 463.37 | 48.51 | 474.83 | 55.33 | -2.38 | 0.02<sup>†</sup> |
| Ventricle volume (cm<sup>3</sup>) |  | 18.04 | 8.16 | 15.01 | 5.38 | 1.68 | - |
| GAF |  | 59.43 | 11.11 | 83.40 | 4.62 | -11.19 | <0.0001<sup>†</sup> |
| PANSS-total |  | 62.07 | 16.71 |  |  |  |  |
| Age at Onset (years) |  | 22.55 | 5.22 |  |  |  |  |
| Duration of illness (days) |  | 691.77 | 511.49 |  |  |  |  |
| CPZ equivalent (mg/day) |  | 340.59 | 272.49 |  |  |  |  |
|  |  | N | % | N | % |  |  |
| Sex | Female | 10 | 33.3 | 17 | 34.7 |  |  |
|  | Male | 20 | 66.7 | 32 | 65.3 |  |  |

### MRI acquisition

Data acquisition was performed as described in our previous works^8,59^. Briefly, MRI scanning sessions were performed on a 3-Tesla system (Magnetom TrioTim, Siemens Healthineers, Erlangen, Germany), equipped with a 32-channel head coil. A 1-mm isotropic T^1^-weighted image was acquired. Whole-brain diffusion-weighted images (DWI) were acquired using diffusion spectrum imaging (DSI) scheme across 15 b-values, ranging from 0 to 8000 s/mm^2^, spatial resolution of 2.2 × 2.2 × 3 mm^3^. Additional acquisition details can be found in the Supplementary Material.

### Image preprocessing

The T_1_-weighted images were bias field corrected^60^, volumes were estimated with FreeSurfer^61^ and normalized by the estimated total intracranial volume (eTICV). The diffusion preprocessing pipeline included MP-PCA denoising and corrections for Gibbs ringing-, EPI distortions-, eddy currents and motion, following most recent guidelines^62^ - see Supplementary Material for preprocessing details.

### Microstructure estimation

For DKI and WMTI-W estimation, the diffusion dataset was truncated^63^ at b≤2500 s/mm^2^. DKI was fit voxel-wise in the entire brain^64^ using a weighted linear-least squares algorithm in Matlab^64^, from which seven scalar maps were derived – four from DTI: radial, mean, axial diffusivity (RD, MD, AD) and FA, and three from DKI: radial, mean, axial kurtosis (RK, MK, AK). WMTI-W parameters were estimated voxel-wise from the seven DTI/DKI scalars, using an in-house Python script (*github*.*com/Mic-map/WMTI-Watson_Python*), yielding five parameter maps: axonal water fraction *f*, intra-axonal diffusivity *D*_*a*_, extra-axonal parallel and perpendicular diffusivities *D*_*e*,||_, *D*_*e*,⊥_ and axon orientation alignment *c*_2_. For each subject, voxels with unphysical values (RD, MD, AD > 4 µm^2^/ms; FA > 1, RK, MK, AK > 10) or negative values in any of the parameters were excluded from the analysis across all parametric maps, if present.

### ^1^H Magnetic resonance spectroscopy

All MRS measurements were performed on a 3T Trio MR scanner (Siemens Medical Solutions, Erlangen, Germany) with a TEM volume coil. B_0_ field inhomogeneity was optimized using 1^st^ and 2^nd^ order shimming with FAST(EST)MAP^65^. 1H MR spectra were obtained in the voxel located in the mPFC using the SPECIAL^66^ localization sequence (TE/TR=6/4000ms, VOI=20×20×25mm^3^, 148 averages). Water unsuppressed spectra were acquired with the same parameters (2 averages) as an internal reference.

### Spectral quantification

Metabolite concentrations were quantified with LCModel^67^ using unsuppressed water MR spectra as an internal reference. Tissue composition within the MRS voxel was evaluated based on the segmentation of 3D MPRAGE images. Fractions of white matter (WM), gray matter (GM) and cerebrospinal fluid (CSF) were used to correct for water content in the measurement voxel. The GSH and Glu levels were reported previously^68^.

### Statistical analysis

#### Tract-based spatial statistic

Associations between MRS metabolite concentrations and dMRI microstructure metrics were tested at voxel level using FSL’s Tract-Based Spatial Statistics^69^ (TBSS). First, individual FA maps were used to build a study-specific FA template using ANTs^70^ and the estimated warps were applied to each of the 12 dMRI parametric maps for spatial normalization. From the average FA, the WM skeleton mask (FA>0.25) was estimated and used for permutation testing (FSL randomise^71^, 5000 permutations) on all dMRI metrics. The resulting statistical maps were corrected using False-Discovery Rate (FDR) and Threshold-Free Cluster Enhancement (TFCE). Significant voxels were averaged and plotted alongside the cluster maps for each significant contrast. Additionally, we report only the associations with average cluster p-value<0.0083 (0.05/6 contrasts). The relationship between metabolite concentration and each microstructure metric was modeled as

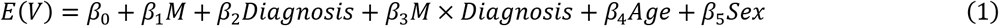

where *E*(*V*) is the expected value of a voxel, and *M* is one of the metabolites of interest. A correction for age and sex was included systematically.

#### Post hoc analyses

To verify if the other metabolites could contribute or suppress the associations within the myo-Ins clusters, we re-computed the association between myo-Ins and each microstructure metric in the significant clusters combined, identified with TBSS at the previous step, by regressing myo-Ins together with the concentrations of the other metabolites:

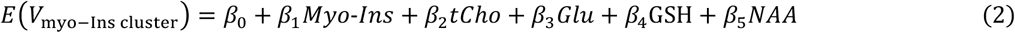

Finally, we investigated whether myo-Ins is influenced by treatment, illness duration, or substance use (e.g., cannabis) which could influence metabolite levels or their association with WM microstructure, given their known impact on brain metabolism^72,73^. For details see Post Hoc Analyses Methodology in the Supplementary Material.

## Results

### Demographics

Summary demographics of the cohort can be found in **Table 1**. EP (24±6 y/o) and HC (25±6 y/o) did not differ in age (p=0.40). EP participants showed lower normalized total brain (p=0.0068) volume and lower normalized white matter (p=0.020) volume than HC. Functioning levels as measured by the GAF were lower in EP than HC (p<0.0001). Noteworthy is an imbalance between males (~66%) and females (~34%) among both EP and HC participants.

### Group differences in metabolites and WM microstructure

We did not find any difference between EP and HC in any metabolite concentrations (p>0.17). Total Choline (p<0.0001) and NAA (p=0.0024) were positively associated with age but only tCho showed differences between EP and HC in the age effect (age × diagnosis interaction, p=0.017). Metabolite concentrations were not associated with the current dose of antipsychotic medication (chlorpromazine equivalent, p>0.12).

Differences between EP and HC in the WM microstructure were reported in our previous work in a larger cohort of which the current one is a subset (allowing for availability of both dMRI and MRS data in the same participant)^8^. Briefly, we found increased DTI diffusivities and reduced kurtosis in EP vs HC (mean |*Cohen’s d*|=0.46). WMTI-W attributed the findings primarily to reduced axonal water fraction, *f*, and changes in the extra-axonal compartment, evident from the higher extra-axonal parallel and perpendicular diffusivities *D*_*e*,||_, *D*_*e*,⊥_ in EP compared to HC. In the current smaller cohort subset we only found significantly higher MD, AD, and *D*_*e*,||_ (p<0.035).

### Associations between myo-inositol and total choline concentrations and WM microstructure

Several clusters of associations were found in WM. Overall, we found that myo-Ins showed the largest clusters of association with the WMTI-W metrics, but not with DTI or DKI metrics (except for axial diffusivity, AD). No associations with tCho were found.

#### Myo-Inositol

Myo-Ins showed the widest and strongest association with dMRI metrics in EP. WMTI-W revealed a positive association of myo-Ins with the axonal water fraction, *f* (p<0.0001, **Fig. 1A**), and a negative association with the extra-axonal parallel diffusivity, *D*_*e*,||_ (p<0.0001, **Fig. 1C**) in EP, with significantly different slopes from HC (*f, D*_*e*,||_: p<0.0001, **Fig. 1B, D**). Clusters were located primarily in the forceps minor (**Fig. 1C**), genu and body of the corpus callosum (**Fig. 1**; Hofer and Frahm^74^: region *I*-prefrontal, *II*-premotor and supplementary motor, and *III*-motor), and to a lesser extent the anterior corona radiata (**Fig. 1A**). Myo-Ins was also negatively associated with AD (p<0.0001, **Fig. S1**), but only in the left internal capsule.

**Figure 1.**
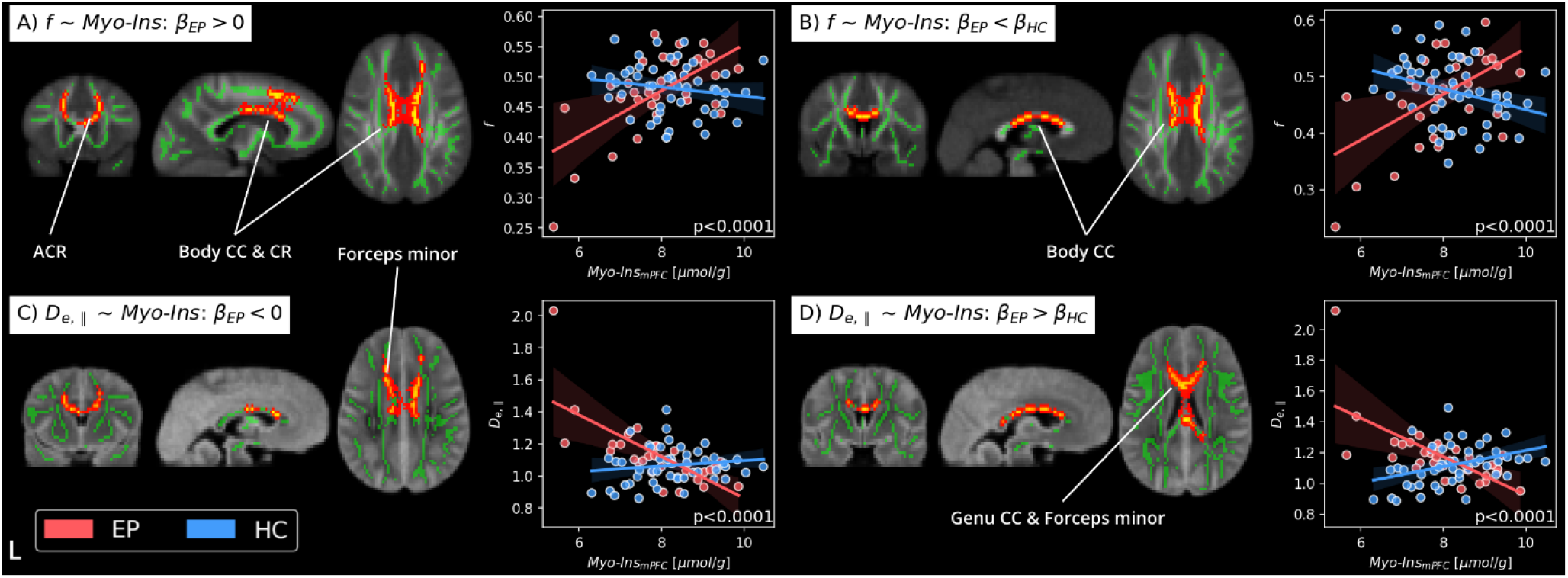
TBSS clusters of associations between *f* and *D*_*e*,||_ and myo-inositol concentration in the mPFC. Centered around the genu, body of the corpus callosum (CC) and some parts of the anterior corona radiata (ACR) and the forceps minor, WMTI-W identifies lower axonal water fraction, *f* (A), and higher extra-axonal parallel diffusivity, *D*_*e*,||_ (C) with lower myo-Ins concentration. In HC, no relationship between myo-Ins and dMRI metrics in the WM was found, while the slopes of EP significantly differed from HC for all the metrics in the same areas (B, D). Note, removing the datapoints with more extreme values (i.e. *f*<0.35 and *D*_*e*,||_>1.6) does not change the results. *β<SUB>EP</SUB>>0*: contrast testing the slope of EP is significantly positive or negative. *β<SUB>EP</SUB>> β<SUB>HC</SUB>*: contrast testing the difference in slopes between EP and HC is significant, L: left hemisphere.

**Figure 2.**
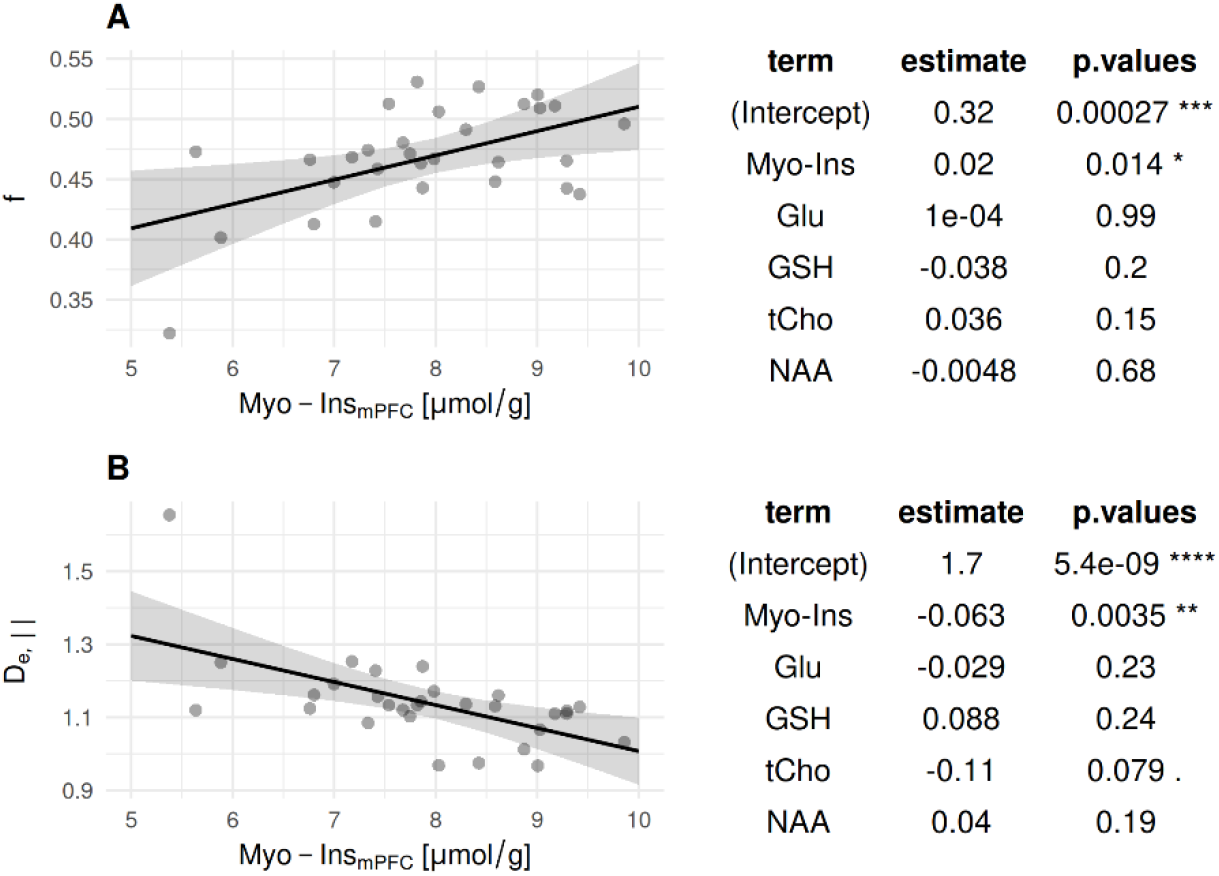
Association of EP’s WM microstructure values in the post hoc cluster with all the metabolite concentrations (myo-Ins, Glu, GSH, tCho, and NAA, see **Eq. 2**). Myo-Inositol drives most of the associations with the WM metrics and is not suppressed by including the other metabolites. Adjusted-R^2^: *f*=0.23 and *D*_*e*,||_=0.36.

### Post Hoc Analysis

When investigating the association of the other metabolites (NAA, GSH, Glu), only one small cluster of WM microstructure association with NAA concentration was found. Higher NAA levels were associated with lower *D*_*a*_ (p<0.0001) in parts of the left posterior and superior corona radiata (**Fig. S2**). We did not find associations with GSH, or Glu.

The frequency mask of the combined WM clusters associated with myo-Ins concentration (**Fig. S3**) showed spatial proximity to the MRS voxel in the mPFC, indicating regional specificity of WM microstructure changes to myo-Ins levels. When the EP’s average cluster mask values were regressed with all metabolites together, the myo-Ins association was not suppressed, confirming the specificity of myo-Ins concentration correlating with WM microstructure in EP: in the *f* regression model (**Fig. 3A**), myo-Ins was the sole significant metabolite (p<0.014, *f* adjusted-R^2^=0.23). In the *D*_*e*,||_ model (**Fig. 3B**), myo-Ins always drove the association (p<0.0035) but tCho showed a trend (p=0.079, *D*_*e*,||_ adjusted-R^2^=0.36).

Furthermore, to ensure robustness of our findings, we repeated the post hoc analyses excluding the EP participant with the most extreme value (*f*<0.35 and *D*_*e*,||_>1.6 µm^2^/ms), but it did not change the results (p<0.028 for *f* and *D*_*e*,||_ slope and group interactions). Excluding female EP participants (who are a minority) also did not change the myo-Ins association with dMRI metrics (p<0.039 for *f* and *D*_*e*,||_ slope and group interactions).

The results for the analyses including medication, drugs or cannabis exposure are reported in the Post Hoc Analyses Sections in the Supplementary Material. In short, none of the confounding variables influenced the relationship of WM microstructure (*f, D*_*e*,||_) with myo-Ins.

## Discussion

In this work, we linked the mPFC myo-Ins and tCho concentrations, which are glial markers^35^, to the WM microstructure features in EP. Myo-Ins concentration in the mPFC showed the strongest and largest association clusters to the WM microstructure among all investigated metabolites. In EP, myo-Ins correlated positively with axonal water fraction, *f*, while large clusters of negative associations were also found with *D*_*e*,||_. However, we did not find such associations in HC nor with tCho.

In an exploratory fashion, we also investigated associations with three major metabolites known to be involved in schizophrenia, Glu^20^, GSH^19^ and NAA^55^, but we found limited associations with the microstructure, except for a negative association between NAA concentration and *D*_*a*_ in EP, confirming myo-Ins as the most prominent metabolite associated with WM alterations in EP. Finally, we verified that our findings were not influenced by indirect effects of treatment, substance exposure^72,73^ or other demographic characteristics.

Microstructure metrics are considered sensitive to myelination but also to glial cell density^11^ as increases or decreases in tissue cellularity could hinder or facilitate water movement, respectively, altering the diffusivity properties of the WM^11,75^. In cuprizone-intoxicated mice^75^, the initial acute inflammation sharply lowers extra-axonal diffusivities (↓*D*_*e*,||_, ↓*D*_*e*,⊥_) due to cellular crowding of the extra-axonal space as a result of gliosis. Once acute inflammation subsides, demyelination dominates, reducing the axonal water fraction (↓*f*) while raising diffusivities (↑*D*_*e*,||_, ↑*D*_*e*,⊥_), relative to controls. Furthermore, longitudinal validation studies related reduced parallel diffusivity to astrogliosis^75–77^, and a diffusion-weighted MRS study using the same model of cuprizone intoxication showed that myo-Ins diffusivity rises with histologically-confirmed astrocyte hypertrophy^78^. Together, these findings suggest that astrocytic changes directly influence diffusivity metrics.

Contrary to our expectations, the average myo-Ins concentrations in the EP group did not differ from HC in our cohort. In their review, Das et al. reported that in studies with higher number of female patients, the myo-inositol reduction is much more pronounced^41^. Thus, the absence of such group differences in metabolite concentrations in our study may be due to the sex imbalance in our EP cohort (20 male vs 10 female), but our power to test sex × diagnosis interactions was limited.

However, the TBSS analysis (corrected for age and sex) showed that participants with the most altered WM (estimates further away from HC, **Fig. 1A,C**) had the lowest mPFC myo-Ins levels. Low myo-Ins may indicate a reduction in astrocyte density, integrity (e.g. atrophy^26,79^) or deficits in astrocyte activation and recruitment^41^ in the mPFC, which may extend beyond that area to neighboring WM tracts. The main clusters of associations included consistently the anterior corona radiata, forceps minor, and the genu and body of the corpus callosum, all white matter regions and fiber bundles adjacent or with projection to and from the mPFC, the target area of the MRS voxel. This suggests that mPFC myo-Ins concentrations are related to the WM microstructure both locally (within the MRS voxel: forceps minor) and proximally (adjacent or connected to the MRS voxel: corpus callosum body and corona radiata). Notably, the anterior corona radiata and corpus callosum have consistently been identified as among the most affected regions in SZ^3,8^ and early-onset SZ^80^.

Relative to distribution values in HC, only a subset of EP patients displays lower myo-Ins, lower *f* and higher *D*_*e*,||_ which may constitute a patient subgroup. In a meta-analysis of Das et al.^41^, the authors reported a small but statistically significant reduction in medial prefrontal myo-inositol levels (effect size = 0.2) in individuals with SZ. In the light of the small effect size, the authors proposed that the biological pathways affecting the myo-inositol and astroglia were likely to operate only in a subset of patients with schizophrenia^41^. Remarkably, our findings strongly support this interpretation, suggesting that a combination of MRS mPFC myo-Ins and WMTI-W metrics could serve as a potential imaging biomarker for stratifying patients into subgroups. Similarly, glial changes are a core feature of SZ^26^. Electron microscopy studies showed dystrophic and swollen astrocytes in SZ^81^. However, GFAP-based and Nissl staining studies reported more controversial findings, indicating that the density reduction may not be extensive^26^, or possibly characterize only a subset of patients^41^ which seems to be the case in our cohort. For example, in their meta-analysis Trépanier et al.^82^ found no consistent alterations in glial fibrillary acidic protein (GFAP) expression, a result potentially explained by the heterogeneity of the SZ population, which may comprise distinct subgroups with and without astrocytic abnormalities.^3,8,80^

The causal relation between mPFC myo-Ins and WMTI-W *f* and *D*_*e*,||_ is complex and merits dedicated investigation beyond the current work. Here, we limit ourselves to some possible explanations: an astrocyte dysfunction more broadly affecting and propagating to the frontal WM in a subset of EP may be an interpretation for the observed association patterns. A reduction in the water hindrance due to the less dense astrocyte crowding of the WM extra-axonal space would be captured as a decrease in (↓*f*), and an increase of extra-axonal parallel diffusivity (↑*D*_*e*,||_)^11,75–77^. This would also suggest that the WMTI-W parameters *f* and *D*_*e*,||_ may have higher specificity to WM alterations caused by glial changes than DKI and DTI scalars. In parallel, myo-Ins has an osmolytic^36,83^ effect (i.e. regulates cellular osmotic pressure), thus low myo-Ins may also be related to astrocyte and glia volume reduction or atrophy^26^ in the WM, which has been found in the prefrontal WM of SZ speciments^26,79^, and could show similar WM patterns (↓*f*, ↑*D*_*e*,||_) at low myo-Ins concentrations. Alternatively, reduced oligodendrocyte density of up to 30% is a common replicated finding in SZ^26,84,85^. These cells provide metabolic^86^ and myelination support^87^ to the neurons. A reduced number of oligodendrocytes would yield a similar pattern to astrocytic deficits, via affected myelination and increased extra-axonal mobility (↓*f*, ↑*D*_*e*,||_, ↑*D*_*e,⊥*_,), which is consistent with our findings in the current work and in our previous work^8^.

Myo-Ins levels correlating primarily with *D*_*e*,||_ rather than intra-axonal diffusivity (*D*_*a*_) further supports the extra-axonal nature of these changes, corroborating our previous reports of altered extra-axonal compartment in EP and SZ^88^. Notably, the lack of correlations in the same regions for Glu and NAA, intraneural metabolites^48,56^, reinforces this interpretation.

We also reported a negative association between AD and myo-Ins in the internal capsule. Such association may reflect the indirect effect of myo-Ins changes to the mPFC axons projecting to and from the internal capsule.

To our knowledge, this is the second work investigating the relationship of WM microstructure with myo-Ins and tCho in schizophrenia, and the first one reporting on EP together with specific WM microstructure metrics that aid the biological interpretation. In the work of Chiappelli and colleagues^89^, the authors reported a negative association between FA and myo-Ins in both SZ and HC, and concluded that, among other metabolites such as NAA, Glu and tCho, myo-Ins was the only one driving the FA changes. The negative association with FA was interpreted as evidence of an effect of inflammation on WM microstructure. Here, we did not find any association with FA and the significant associations were only in the EP group, but not in the HC. Several factors may explain this discrepancy. First, we measured the myo-Ins concentration in the frontal GM dominated area and not purely in the WM. Second, our cohort exclusively included EP instead of SZ. Third, our participants were younger (average age of ~25 vs ~39 years old) with a narrower age distribution (18-36 vs 20-58 years old), although Chiappelli et al. reported no age effect on such association.

Aside from myo-Ins, the sole other metabolite we found associated with WM microstructure was NAA. In EP, NAA was found to be negatively associated with *D*_*a*_ in parts of the left superior and posterior corona radiata, so in different regions than for myo-Ins, possibly indicating an indirect relation between mPFC NAA and less complex and ramified intra-axonal space, axonal metabolism^90^, or NAA diffusion^91^ in the corona radiata of EP.

The lack of association between WM and tCho, Glu, or GSH does not exclude their role in SZ but suggests that they may not directly contribute to WM alteration in EP, or that our study lacks sufficient power to detect their effect. This view is supported by the age-tCho relationship and its trend difference between EP and HC, which could act as a confounder, misattributing covariance between metabolites and WM to age or vice versa.

The absence of associations with GSH is in line with our previous work on the peripheral GSH-redox system^92^. However, we did not replicate the previously reported association between GSH levels and the average generalized FA of the cingulum WM bundle^93^, possibly due to cohort differences (EP vs SZ). Taken together, this suggests that the effect of GSH on white matter may be subtle, potentially requiring larger regions of interest to detect it. As such, the TBSS method may lack the sensitivity to capture this effect.

As last set of analyses, we also tested the effects of treatment, illness duration, and substance exposure^72,73^ on our results, to verify our findings were not influenced or caused by unaccounted confounding effects. Previous works showed that lower myo-Ins was related to the severity of drug and cannabis abuse^72^, while myo-Ins levels were higher in treatment-naїve first-episode psychosis than controls^44^, and higher in treatment resistant than non-treatment resistant SZ or HC^45^. However, we found no association with antipsychotic medication, illness duration or substance exposure, indicating that the links between myo-Ins concentrations and WM microstructure in neighboring tracts are independent of these factors.

### Limitations

The first limitation of our work is the different spatial localization of the MRS and dMRI signals. Whereas the dMRI metrics were computed and tested in the WM, our single voxel MRS acquisition was centered on the mPFC and contained primarily GM together with some WM and CSF fractions. However, the significant clusters were spatially coherent with the location of the MRS voxel, supporting a plausible microstructural–metabolic WM-GM coupling. Additionally, by employing a spatially unbiased approach such as TBSS rather than a region-of-interest method, we identified anatomically meaningful clusters aligned with the MRS voxel placement in a data-driven way, which strengthens the interpretability and validity of our findings. Furthermore, the sample size of our EP cohort is relatively small (n=30) and featured more males (n=20) than females (n=10). However, excluding the female group did not change the substance of our findings. Despite a systematic correction for age and sex in our tests, and the narrow age span of our participants, we cannot fully exclude the influence of maturation and aging on the WM microstructure and metabolite concentrations (myo-Ins and tCho increase, Glu and NAA decrease with aging^94^). Finally, despite our best efforts, the measures used for medication (chlorpromazine-equivalent dose at the time of the scan) and substance use are imperfect and may not capture the patients’ cumulative drugs histories and effects.

## Conclusions

In this work we showed for the first time the comprehensive associations between mPFC concentrations of glial metabolites and advanced dMRI WM microstructure metrics in EP. We demonstrated that lower levels of mPFC myo-inositol in patients are strongly associated with proximal changes in WM tissue diffusivities that corresponded to lower axonal water fraction and less hindered extra-axonal parallel diffusivity (higher *D*_*e*,||_, lower *f*), suggesting white matter alteration could be linked to astrocytic changes in a subgroup of early psychosis patients. Furthermore, joint assessment of myo-inositol levels and white matter microstructure metrics may serve as a potential imaging biomarker for stratifying patients into subgroups, possibly characterized by glia-driven pathophysiology.

## Data Availability

All data produced in the present study are available upon reasonable request to the authors

## Acknowledgments

This work was supported by the Swiss National Science Foundation (PCEFP2_194260, to I.J.; 320030_197787 to PH and YA), the National Center of Competence in Research (NCCR) “SYNAPSY - The Synaptic Bases of Mental Diseases” from the Swiss National Science Foundation (n° 51NF40 – 185897 to KQD & PC) and the Foundation Alamaya. Dr. Alameda is supported by Carigest fellowship and by Frutiger Adrian et Simone fellowship. Dr. Dwir and P. Klauser are supported by Frutiger Adrian & Simone fellowship. Dr. L. Xin is supported by SNSF Consolidator grant n° 213769. We acknowledge the resources and expertise provided by the CIBM Center for Biomedical Imaging.

## References

1. Wittchen HU, Jacobi F, Rehm J, Gustavsson A, Svensson M, Jönsson B, et al. The size and burden of mental disorders and other disorders of the brain in Europe 2010. Eur Neuropsychopharmacol J Eur Coll Neuropsychopharmacol. 2011;21(9):655–679. doi:10.1016/j.euroneuro.2011.07.018

2. Di Biase MA, Pantelis C, Zalesky A. White Matter Pathology in Schizophrenia. In: Kubicki M, Shenton ME, eds. Neuroimaging in Schizophrenia. Cham: Springer International Publishing; 2020:71–91. doi:10.1007/978-3-030-35206-6_4

3. Kelly S, Jahanshad N, Zalesky A, Kochunov P, Agartz I, Alloza C, et al. Widespread white matter microstructural differences in schizophrenia across 4322 individuals: results from the ENIGMA Schizophrenia DTI Working Group. Mol Psychiatry. 2018;23(5):1261–1269. doi:10.1038/mp.2017.170

4. Beaulieu C. The basis of anisotropic water diffusion in the nervous system - a technical review. NMR Biomed. 2002;15(7-8):435-455. doi:10.1002/nbm.782

5. Waszczuk K, Tyburski E, Rek-Owodziń K, Plichta P, Rudkowski K, Podwalski P, et al. Relationship between White Matter Alterations and Pathophysiological Symptoms in Patients with Ultra-High Risk of Psychosis, First-Episode, and Chronic Schizophrenia. Brain Sci. 2022;12(3):354. doi:10.3390/brainsci12030354

6. Sagarwala R, Nasrallah HA. The effect of antipsychotic medications on white matter integrity in first-episode drug-naïve patients with psychosis: A review of DTI studies. Asian J Psychiatry. 2021;61:102688. doi:10.1016/j.ajp.2021.102688

7. Klauser P, Baker ST, Cropley VL, Bousman C, Fornito A, Cocchi L, et al. White Matter Disruptions in Schizophrenia Are Spatially Widespread and Topologically Converge on Brain Network Hubs. Schizophr Bull. 2017;43(2):425–435. doi:10.1093/schbul/sbw100

8. Pavan T, Alemán-Gómez Y, Jenni R, Steullet P, Schilliger Z, Dwir D, et al. White matter microstructure alterations in early psychosis and schizophrenia. Transl Psychiatry. 2025;15(1):1–10. doi:10.1038/s41398-025-03397-1

9. Jensen JH, Helpern JA. MRI quantification of non-Gaussian water diffusion by kurtosis analysis. NMR Biomed. 2010;23(7):698–710. doi:10.1002/nbm.1518

10. Jespersen SN, Olesen JL, Hansen B, Shemesh N. Diffusion time dependence of microstructural parameters in fixed spinal cord. NeuroImage. 2018;182:329–342. doi:10.1016/j.neuroimage.2017.08.039

11. Jelescu IO, Fieremans E. Chapter 2 - Sensitivity and specificity of diffusion MRI to neuroinflammatory processes. In: Laule C, Port JD, eds. Advances in Magnetic Resonance Technology and Applications. Vol 9. Imaging Neuroinflammation. Academic Press; 2023:31–50. doi:10.1016/B978-0-323-91771-1.00010-1

12. Jelescu IO, Budde MD. Design and Validation of Diffusion MRI Models of White Matter. Front Phys. 2017;5. https://www.frontiersin.org/articles/10.3389/fphy.2017.00061. xAccessed December 1, 2022.

13. Fieremans E, Jensen JH, Helpern JA. White matter characterization with diffusional kurtosis imaging. NeuroImage. 2011;58(1):177–188. doi:10.1016/j.neuroimage.2011.06.006

14. Di Biase MA, Cetin-Karayumak S, Lyall AE, Zalesky A, Cho KIK, Zhang F, et al. White matter changes in psychosis risk relate to development and are not impacted by the transition to psychosis. Mol Psychiatry. 2021;26(11):6833–6844. doi:10.1038/s41380-021-01128-8

15. Carletti F, Woolley JB, Bhattacharyya S, Perez-Iglesias R, Fusar Poli P, Valmaggia L, et al. Alterations in White Matter Evident Before the Onset of Psychosis. Schizophr Bull. 2012;38(6):1170–1179. doi:10.1093/schbul/sbs053

16. Mittal VA, Dean DJ, Bernard JA, Orr JM, Pelletier-Baldelli A, Carol EE, et al. Neurological Soft Signs Predict Abnormal Cerebellar-Thalamic Tract Development and Negative Symptoms in Adolescents at High Risk for Psychosis: A Longitudinal Perspective. Schizophr Bull. 2014;40(6):1204–1215. doi:10.1093/schbul/sbt199

17. Alba-Ferrara LM, de Erausquin GA. What does anisotropy measure? Insights from increased and decreased anisotropy in selective fiber tracts in schizophrenia. Front Integr Neurosci. 2013;7:9. doi:10.3389/fnint.2013.00009

18. Bakhshi K, Chance SA. The neuropathology of schizophrenia: A selective review of past studies and emerging themes in brain structure and cytoarchitecture. Neuroscience. 2015;303:82–102. doi:10.1016/j.neuroscience.2015.06.028

19. Cuenod M, Steullet P, Cabungcal JH, Dwir D, Khadimallah I, Klauser P, et al. Caught in vicious circles: a perspective on dynamic feed-forward loops driving oxidative stress in schizophrenia. Mol Psychiatry. November 2021. doi:10.1038/s41380-021-01374-w

20. McCutcheon RA, Krystal JH, Howes OD. Dopamine and glutamate in schizophrenia: biology, symptoms and treatment. World Psychiatry. 2020;19(1):15–33. doi:10.1002/wps.20693

21. Hu W, MacDonald ML, Elswick DE, Sweet RA. The glutamate hypothesis of schizophrenia: evidence from human brain tissue studies. Ann N Y Acad Sci. 2015;1338(1):38–57. doi:10.1111/nyas.12547

22. Na KS, Jung HY, Kim YK. The role of pro-inflammatory cytokines in the neuroinflammation and neurogenesis of schizophrenia. Prog Neuropsychopharmacol Biol Psychiatry. 2014;48:277–286. doi:10.1016/j.pnpbp.2012.10.022

23. Comer AL, Carrier M, Tremblay MEÈ, Cruz-Martín A. The Inflamed Brain in Schizophrenia: The Convergence of Genetic and Environmental Risk Factors That Lead to Uncontrolled Neuroinflammation. Front Cell Neurosci. 2020;14. doi:10.3389/fncel.2020.00274

24. Bishop JR, Zhang L, Lizano P. Inflammation Subtypes and Translating Inflammation-Related Genetic Findings in Schizophrenia and Related Psychoses: A Perspective on Pathways for Treatment Stratification and Novel Therapies. Harv Rev Psychiatry. 2022;30(1):59. doi:10.1097/HRP.0000000000000321

25. Cassoli JS, Guest PC, Malchow B, Schmitt A, Falkai P, Martins-de-Souza D. Disturbed macro-connectivity in schizophrenia linked to oligodendrocyte dysfunction: from structural findings to molecules. NPJ Schizophr. 2015;1:15034. doi:10.1038/npjschz.2015.34

26. Bernstein HG, Nussbaumer M, Vasilevska V, Dobrowolny H, Nickl-Jockschat T, Guest PC, et al. Glial cell deficits are a key feature of schizophrenia: implications for neuronal circuit maintenance and histological differentiation from classical neurodegeneration. Mol Psychiatry. December 2024:1–15. doi:10.1038/s41380-024-02861-6

27. Liu SH, D. Y, Chen L, Cheng Y. Glial Cell Abnormalities in Major Psychiatric Diseases: A Systematic Review of Postmortem Brain Studies. Mol Neurobiol. 2022;59(3):1665–1692. doi:10.1007/s12035-021-02672-8

28. Hof PR, Haroutunian V, Copland C, Davis KL, Buxbaum JD. Molecular and cellular evidence for an oligodendrocyte abnormality in schizophrenia. Neurochem Res. 2002;27(10):1193–1200. doi:10.1023/a:1020981510759

29. Hof PR, Haroutunian V, Friedrich VL, Byne W, Buitron C, Perl DP, et al. Loss and altered spatial distribution of oligodendrocytes in the superior frontal gyrus in schizophrenia. Biol Psychiatry. 2003;53(12):1075–1085. doi:10.1016/S0006-3223(03)00237-3

30. Volk DW, Moroco AE, Roman KM, Edelson JR, Lewis DA. The Role of the Nuclear Factor-κB Transcriptional Complex in Cortical Immune Activation in Schizophrenia. Biol Psychiatry. 2019;85(1):25–34. doi:10.1016/j.biopsych.2018.06.015

31. Catts VS, Wong J, Fillman SG, Fung SJ, Shannon Weickert C. Increased expression of astrocyte markers in schizophrenia: Association with neuroinflammation. Aust N Z J Psychiatry. 2014;48(8):722–734. doi:10.1177/0004867414531078

32. Zhang Y, Catts VS, Sheedy D, McCrossin T, Kril JJ, Shannon Weickert C. Cortical grey matter volume reduction in people with schizophrenia is associated with neuro-inflammation. Transl Psychiatry. 2016;6(12):e982. doi:10.1038/tp.2016.238

33. Pasternak O, Sochen N, Gur Y, Intrator N, Assaf Y. Free water elimination and mapping from diffusion MRI. Magn Reson Med. 2009;62(3):717–730. doi:10.1002/mrm.22055

34. Di Biase MA, Zalesky A, Cetin-Karayumak S, Rathi Y, Lv J, Boerrigter D, et al. Large-Scale Evidence for an Association Between Peripheral Inflammation and White Matter Free Water in Schizophrenia and Healthy Individuals. Schizophr Bull. 2021;47(2):542–551. doi:10.1093/schbul/sbaa134

35. Wijtenburg SA, Rowland LM. Chapter 19 - Schizophrenia spectrum disorders. In: Laule C, Port JD, eds. Advances in Magnetic Resonance Technology and Applications. Vol 9. Imaging Neuroinflammation. Academic Press; 2023:469–487. doi:10.1016/B978-0-323-91771-1.00008-3

36. Rae CD. A Guide to the Metabolic Pathways and Function of Metabolites Observed in Human Brain 1H Magnetic Resonance Spectra. Neurochem Res. 2014;39(1):1–36. doi:10.1007/s11064-013-1199-5

37. Glanville NT, Byers DM, Cook HW, Spence MW, Palmer FBStC. Differences in the metabolism of inositol and phosphoinositides by cultured cells of neuronal and glial origin. Biochim Biophys Acta BBA - Lipids Lipid Metab. 1989;1004(2):169–179. doi:10.1016/0005-2760(89)90265-8

38. Hattingen E, Raab P, Franz K, Zanella FE, Lanfermann H, Pilatus U. Myo-Inositol: a marker of reactive astrogliosis in glial tumors? NMR Biomed. 2008;21(3):233–241. doi:10.1002/nbm.1186

39. Ligneul C, Palombo M, Hernández-Garzón E, Carrillo-de Sauvage MA, Flament J, Hantraye P, et al. Diffusion-weighted magnetic resonance spectroscopy enables cell-specific monitoring of astrocyte reactivity in vivo. NeuroImage. 2019;191:457–469. doi:10.1016/j.neuroimage.2019.02.046

40. Jeon P, Mackinley M, Théberge J, Palaniyappan L. The trajectory of putative astroglial dysfunction in first episode schizophrenia: a longitudinal 7-Tesla MRS study. Sci Rep. 2021;11(1):22333. doi:10.1038/s41598-021-01773-7

41. Das TK, Dey A, Sabesan P, Javadzadeh A, Théberge J, Radua J, et al. Putative Astroglial Dysfunction in Schizophrenia: A Meta-Analysis of 1H-MRS Studies of Medial Prefrontal Myo-Inositol. Front Psychiatry. 2018;9. doi:10.3389/fpsyt.2018.00438

42. Chiappelli J, Rowland LM, Wijtenburg SA, Muellerklein F, Tagamets M, McMahon RP, et al. Evaluation of Myo-Inositol as a Potential Biomarker for Depression in Schizophrenia. Neuropsychopharmacology. 2015;40(9):2157–2164. doi:10.1038/npp.2015.57

43. Chhetri DR. Myo-Inositol and Its Derivatives: Their Emerging Role in the Treatment of Human Diseases. Front Pharmacol. 2019;10:1172. doi:10.3389/fphar.2019.01172

44. Plitman E, de la Fuente-Sandoval C, Reyes-Madrigal F, Chavez S, Gómez-Cruz G, León-Ortiz P, et al. Elevated Myo-Inositol, Choline, and Glutamate Levels in the Associative Striatum of Antipsychotic-Naive Patients With First-Episode Psychosis: A Proton Magnetic Resonance Spectroscopy Study With Implications for Glial Dysfunction. Schizophr Bull. 2016;42(2):415–424. doi:10.1093/schbul/sbv118

45. Smucny J, Carter CS, Maddock RJ. Greater Choline-Containing Compounds and Myo-inositol in Treatment-Resistant Versus Responsive Schizophrenia: A 1H-Magnetic Resonance Spectroscopy Meta-analysis. Biol Psychiatry Cogn Neurosci Neuroimaging. 2024;9(2):137–145. doi:10.1016/j.bpsc.2023.10.008

46. Herminghaus S, Pilatus U, Möller-Hartmann W, Raab P, Lanfermann H, Schlote W, et al. Increased choline levels coincide with enhanced proliferative activity of human neuroepithelial brain tumors. NMR Biomed. 2002;15(6):385–392. doi:10.1002/nbm.793

47. Gill SS, Small RK, Thomas DGT, Patel P, Porteous R, Van Bruggen N, et al. Brain metabolites as 1H NMR markers of neuronal and glial disorders. NMR Biomed. 1989;2(5-6):196–200. doi:10.1002/nbm.1940020505

48. Urenjak J, Williams SR, Gadian DG, Noble M. Proton nuclear magnetic resonance spectroscopy unambiguously identifies different neural cell types. J Neurosci. 1993;13(3):981–989. doi:10.1523/JNEUROSCI.13-03-00981.1993

49. De Marco R, Ronen I, Branzoli F, Amato ML, Asllani I, Colasanti A, et al. Diffusion-weighted MR spectroscopy (DW-MRS) is sensitive to LPS-induced changes in human glial morphometry: A preliminary study. Brain Behav Immun. 2022;99:256–265. doi:10.1016/j.bbi.2021.10.005

50. Yang YS, Smucny J, Zhang H, Maddock RJ. Meta-analytic evidence of elevated choline, reduced N-acetylaspartate, and normal creatine in schizophrenia and their moderation by measurement quality, echo time, and medication status. NeuroImage Clin. 2023;39:103461. doi:10.1016/j.nicl.2023.103461

51. Smucny J, Carter CS, Maddock RJ. Magnetic resonance spectroscopic evidence of increased choline in the dorsolateral prefrontal and visual cortices in recent onset schizophrenia. Neurosci Lett. 2022;770:136410. doi:10.1016/j.neulet.2021.136410

52. Moffett JR, Ariyannur P, Arun P, Namboodiri AMA. N-Acetylaspartate and N-Acetylaspartylglutamate in Central Nervous System Health and Disease. In: Stagg C, Rothman D, eds. Magnetic Resonance Spectroscopy. San Diego: Academic Press; 2014:71–90. doi:10.1016/B978-0-12-401688-0.00006-9

53. Harris AD, MacMillan EL. Chapter 4 - MRS in neuroinflammation. In: Laule C, Port JD, eds. Advances in Magnetic Resonance Technology and Applications. Vol 9. Imaging Neuroinflammation. Academic Press; 2023:79–116. doi:10.1016/B978-0-323-91771-1.00012-5

54. Whitehurst TS, Osugo M, Townsend L, Shatalina E, Vava R, Onwordi EC, et al. Proton Magnetic Resonance Spectroscopy of N-acetyl Aspartate in Chronic Schizophrenia, First Episode of Psychosis and High-Risk of Psychosis: A Systematic Review and Meta-Analysis. Neurosci Biobehav Rev. 2020;119:255–267. doi:10.1016/j.neubiorev.2020.10.001

55. Roberts D, Rösler L, Wijnen JP, Thakkar KN. Associations between N-Acetylaspartate and white matter integrity in individuals with schizophrenia and unaffected relatives. Psychiatry Res Neuroimaging. 2023;330:111612. doi:10.1016/j.pscychresns.2023.111612

56. Griffin JL, Bollard M, Nicholson JK, Bhakoo K. Spectral profiles of cultured neuronal and glial cells derived from HRMAS (1)H NMR spectroscopy. NMR Biomed. 2002;15(6):375–384. doi:10.1002/nbm.792

57. Yung AR, Yuen HP, McGorry PD, Phillips LJ, Kelly D, Dell’Olio M, et al. Mapping the onset of psychosis: the Comprehensive Assessment of At-Risk Mental States. Aust N Z J Psychiatry. 2005;39(11-12):964–971. doi:10.1080/j.1440-1614.2005.01714.x

58. Baumann PS, Crespi S, Marion-Veyron R, Solida A, Thonney J, Favrod J, et al. Treatment and Early Intervention in Psychosis Program (TIPP-Lausanne): implementation of an early intervention programme for psychosis in Switzerland. Early Interv Psychiatry. 2013;7(3):322–328. doi:10.1111/eip.12037

59. Alemán-Gómez Y, Baumgartner T, Klauser P, Cleusix M, Jenni R, Hagmann P, et al. Multimodal Magnetic Resonance Imaging Depicts Widespread and Subregion Specific Anomalies in the Thalamus of Early-Psychosis and Chronic Schizophrenia Patients. Schizophr Bull. 2023;49(1):196–207. doi:10.1093/schbul/sbac113

60. Tustison NJ, Avants BB, Cook PA, Zheng Y, Egan A, Yushkevich PA, et al. N4ITK: improved N3 bias correction. IEEE Trans Med Imaging. 2010;29(6):1310–1320. doi:10.1109/TMI.2010.2046908

61. Dale AM, Fischl B, Sereno MI. Cortical Surface-Based Analysis: I. Segmentation and Surface Reconstruction. NeuroImage. 1999;9(2):179–194. doi:10.1006/nimg.1998.0395

62. Ades-Aron B, Veraart J, Kochunov P, McGuire S, Sherman P, Kellner E, et al. Evaluation of the accuracy and precision of the diffusion parameter EStImation with Gibbs and NoisE removal pipeline. NeuroImage. 2018;183:532–543. doi:10.1016/j.neuroimage.2018.07.066

63. Jensen JH, Helpern JA, Ramani A, Lu H, Kaczynski K. Diffusional kurtosis imaging: The quantification of non-gaussian water diffusion by means of magnetic resonance imaging. Magn Reson Med. 2005;53(6):1432–1440. doi:10.1002/mrm.20508

64. Veraart J, Sijbers J, Sunaert S, Leemans A, Jeurissen B. Weighted linear least squares estimation of diffusion MRI parameters: Strengths, limitations, and pitfalls. NeuroImage. 2013;81:335–346. doi:10.1016/j.neuroimage.2013.05.028

65. Gruetter R. Automatic, localized in vivo adjustment of all first- and second-order shim coils. Magn Reson Med. 1993;29(6):804–811. doi:10.1002/mrm.1910290613

66. Mekle R, Mlynárik V, Gambarota G, Hergt M, Krueger G, Gruetter R. MR spectroscopy of the human brain with enhanced signal intensity at ultrashort echo times on a clinical platform at 3T and 7T. Magn Reson Med. 2009;61(6):1279–1285. doi:10.1002/mrm.21961

67. Provencher SW. Estimation of metabolite concentrations from localized in vivo proton NMR spectra. Magn Reson Med. 1993;30(6):672–679. doi:10.1002/mrm.1910300604

68. Xin L, Mekle R, Fournier M, Baumann PS, Ferrari C, Alameda L, et al. Genetic Polymorphism Associated Prefrontal Glutathione and Its Coupling With Brain Glutamate and Peripheral Redox Status in Early Psychosis. Schizophr Bull. 2016;42(5):1185–1196. doi:10.1093/schbul/sbw038

69. Smith SM, Jenkinson M, Johansen-Berg H, Rueckert D, Nichols TE, Mackay CE, et al. Tract-based spatial statistics: voxelwise analysis of multi-subject diffusion data. NeuroImage. 2006;31(4):1487–1505. doi:10.1016/j.neuroimage.2006.02.024

70. Avants BB, Epstein CL, Grossman M, Gee JC. Symmetric diffeomorphic image registration with cross-correlation: evaluating automated labeling of elderly and neurodegenerative brain. Med Image Anal. 2008;12(1):26–41. doi:10.1016/j.media.2007.06.004

71. Winkler AM, Ridgway GR, Webster MA, Smith SM, Nichols TE. Permutation inference for the general linear model. NeuroImage. 2014;92:381–397. doi:10.1016/j.neuroimage.2014.01.060

72. Watts JJ, Garani R, Da Silva T, Lalang N, Chavez S, Mizrahi R. Evidence That Cannabis Exposure, Abuse, and Dependence Are Related to Glutamate Metabolism and Glial Function in the Anterior Cingulate Cortex: A 1H-Magnetic Resonance Spectroscopy Study. Front Psychiatry. 2020;11:764. doi:10.3389/fpsyt.2020.00764

73. Rigucci S, Xin L, Klauser P, Baumann PS, Alameda L, Cleusix M, et al. Cannabis use in early psychosis is associated with reduced glutamate levels in the prefrontal cortex. Psychopharmacology (Berl). 2018;235(1):13–22. doi:10.1007/s00213-017-4745-z

74. Hofer S, Frahm J. Topography of the human corpus callosum revisited—Comprehensive fiber tractography using diffusion tensor magnetic resonance imaging. NeuroImage. 2006;32(3):989–994. doi:10.1016/j.neuroimage.2006.05.044

75. Guglielmetti C, Veraart J, Roelant E, Mai Z, Daans J, Van Audekerke J, et al. Diffusion kurtosis imaging probes cortical alterations and white matter pathology following cuprizone induced demyelination and spontaneous remyelination. NeuroImage. 2016;125:363–377. doi:10.1016/j.neuroimage.2015.10.052

76. Falangola MF, Guilfoyle DN, Tabesh A, Hui ES, Nie X, Jensen JH, et al. Histological correlation of diffusional kurtosis and white matter modeling metrics in cuprizone-induced corpus callosum demyelination. NMR Biomed. 2014;27(8):948–957. doi:10.1002/nbm.3140

77. Wang S, Wu EX, Qiu D, Leung LHT, Lau HF, Khong PL. Longitudinal diffusion tensor magnetic resonance imaging study of radiation-induced white matter damage in a rat model. Cancer Res. 2009;69(3):1190–1198. doi:10.1158/0008-5472.CAN-08-2661

78. Genovese G, Palombo M, Santin MD, Valette J, Ligneul C, Aigrot MS, et al. Inflammation-driven glial alterations in the cuprizone mouse model probed with diffusion-weighted magnetic resonance spectroscopy at 11.7 T. NMR Biomed. 2021;34(4):e4480. doi:10.1002/nbm.4480

79. Hercher C, Chopra V, Beasley CL. Evidence for morphological alterations in prefrontal white matter glia in schizophrenia and bipolar disorder. J Psychiatry Neurosci JPN. 2014;39(6):376–385. doi:10.1503/jpn.130277

88. Pavan T, Alemán-Gómez Y, Jenni R, Steullet P, Schilliger Z, Dwir D, et al. White Matter Microstructure Alterations in Early Psychosis and Schizophrenia. April 2024:2024.02.01.24301979. doi:10.1101/2024.02.01.24301979

89. Chiappelli J, Hong LE, Wijtenburg SA, D. X, Gaston F, Kochunov P, et al. Alterations in frontal white matter neurochemistry and microstructure in schizophrenia: implications for neuroinflammation. Transl Psychiatry. 2015;5(4):e548–e548. doi:10.1038/tp.2015.43

90. Moffett JR, Ross B, Arun P, Madhavarao CN, Namboodiri AMA. N-Acetylaspartate in the CNS: from neurodiagnostics to neurobiology. Prog Neurobiol. 2007;81(2):89–131. doi:10.1016/j.pneurobio.2006.12.003

91. Najac C, Branzoli F, Ronen I, Valette J. Brain intracellular metabolites are freely diffusing along cell fibers in grey and white matter, as measured by diffusion-weighted MR spectroscopy in the human brain at 7 T. Brain Struct Funct. 2016;221(3):1245–1254. doi:10.1007/s00429-014-0968-5

92. Pavan T, Steullet P, Alemán-Gómez Y, Jenni R, Schilliger Z, Cleusix M, et al. Associations Between Blood Markers of the GSH Redox cycle and Brain White Matter Microstructure in Psychosis. September 2024:2024.09.02.24312940. doi:10.1101/2024.09.02.24312940

93. Monin A, Baumann PS, Griffa A, Xin L, Mekle R, Fournier M, et al. Glutathione deficit impairs myelin maturation: relevance for white matter integrity in schizophrenia patients. Mol Psychiatry. 2015;20(7):827–838. doi:10.1038/mp.2014.88

94. Cleeland C, Pipingas A, Scholey A, White D. Neurochemical changes in the aging brain: A systematic review. Neurosci Biobehav Rev. 2019;98:306–319. doi:10.1016/j.neubiorev.2019.01.003

