## Supplementary Material for "Myo-Inositol Concentration in the Medial Prefrontal Cortex is Associated with Changes in Brain White Matter Microstructure in Early Psychosis"

August 8, 2025

### Supplementary Material

#### Methods

##### MRI Acquisition details

Magnetization-prepared rapid acquisition gradient echo (MPRAGE): echo time (TE) 2.98ms, repetition time (TR) 2300 ms, inversion time (TI) 900 ms, field of view (FOV) 160 x 240 x 256 mm$^3$ and voxel size 1 x 1 x 1.2 mm$^3$), acquisition time 7 minutes. Pulsed Gradient Spin-echo (PGSE) echo planar imaging (EPI) sequence: acquired in Cartesian q-space coverage totalling 257 DWI volumes. TE = 103 ms, TR = 5.9 s. FOV = 211 x 211 x 114mm$^3$, voxel size = 2.2 x 2.2 x 3mm$^3$, 96x96x38 slices, partial Fourier = 0.75, 1 b=0 acquisition and 256 directions, acquisition time 13 minutes. Any participants with pacemaker, cochlear implant, implant of stimulator or drug pump, glucose sensor, bypass valve or pregnant were not allowed in the MRI scanner and excluded from the study.

##### MRI Preprocessing details

The diffusion preprocessing pipeline included MP-PCA denoising (Veraart et al., 2016) and Gibbs ringing correction (Kellner et al., 2016; Lee et al., 2021). The EPI distortions were corrected using ANTs non-linear registration since no reverse phase encode image or magnetic field map were available. The registration of the DSI b=0 volume to the MPRAGE was constrained only in the direction of the distortion, and the estimated correction was warped back to each DWI volume in native space (Alemán-Gómez et al., 2023; Tax et al., 2022). Thereafter, the distortion-corrected images were further corrected for eddy currents and motion using FSL *eddy* (Andersson and Sotiropoulos, 2016). Since FSL eddy does not support DSI data natively (see *–data_is_shelled* in: https://fsl.fmrib.ox.ac.uk/fsl/fslwiki/eddy/UsersGuide), temporarily merging or directly dropping a subset of volumes based on their b-values in order to simulate shells was necessary to accommodate the algorithm. Namely, volumes b=1500 and b=2000 were merged as b=1750, b=4000 was merged to b=4500 while b=6000 and b=8000 were removed. Right after the eddy execution the merged volumes were split back into their original b-values except for the dropped volumes (for the dMRI metric estimations only the b-values≤2500 were needed). The reduced DWI images featured 57 directions (1:b=0, 6:b=500, 12:b=1000, 8:b1500, 6:b=2000, 24:b=2500). Data quality was assessed by visual inspection.

#### Post hoc analyses methodology

WM post hoc analyses were computed from the frequency mask of the significant myo-Ins clusters (Fig. 3). The mask was generated by combining all the masks of significant WM clusters across dMRI modalities and contrasts of the myo-Ins associations. We verified that the relationships found were not due to the duration of illness, medication, drugs and cannabis use frequency (see next section ”Substance Exposure”, Watts et al., 2020; Rigucci et al., 2018).

#### Substance Exposure

To evaluate the effect of substance abuse, we use the Case Manager Rating Scale (CMRS). CMRS is a 10 item scale indicating the frequency of use of 10 different substances from 1 (absent) to 5 (extremely severe). The substances tracked are: Cannabis, Cocaine, Hallucinogens, Opiates, PCP, Stimulants, Sedatives/Hypnotics/Anxiolytics, Non-prescribed medications, and Others. The cumulative rating is computed by summing all the individual scales and subtracting 9 (so absent=1). Every participant with CMRS≥2 (cannabis and total) was classified as exposed. Among the EP cohort, we found CMRS-Cannabis: non-exposed=14, exposed=9 subjects (mean score=2.44). CMRS-Total: non-exposed=7, exposed=16 subjects (mean score=2.48).

#### Diagnosis

| Diagnosis | | N | % |
|---|---|---|---|
| | Schizophrenia | 16 | 53.3 |
| | Bipolar disorder | 1 | 3.3 |
| | Bipolar disorder with psychotic features | 1 | 3.3 |
| | Depressive disorder with psychotic features | 1 | 3.3 |
| | Brief psychotic disorder | 4 | 13.3 |
| | Unspecified psychotic disorder | 2 | 6.7 |
| | Schizoaffective disorder | 5 | 16.7 |

Table S 1: Early Psychosis patients' individual diagnoses.

### Supplementary results

#### Associations between myo-ins and tCho MRS metabolites and WM microstructure

##### Myo-Inositol

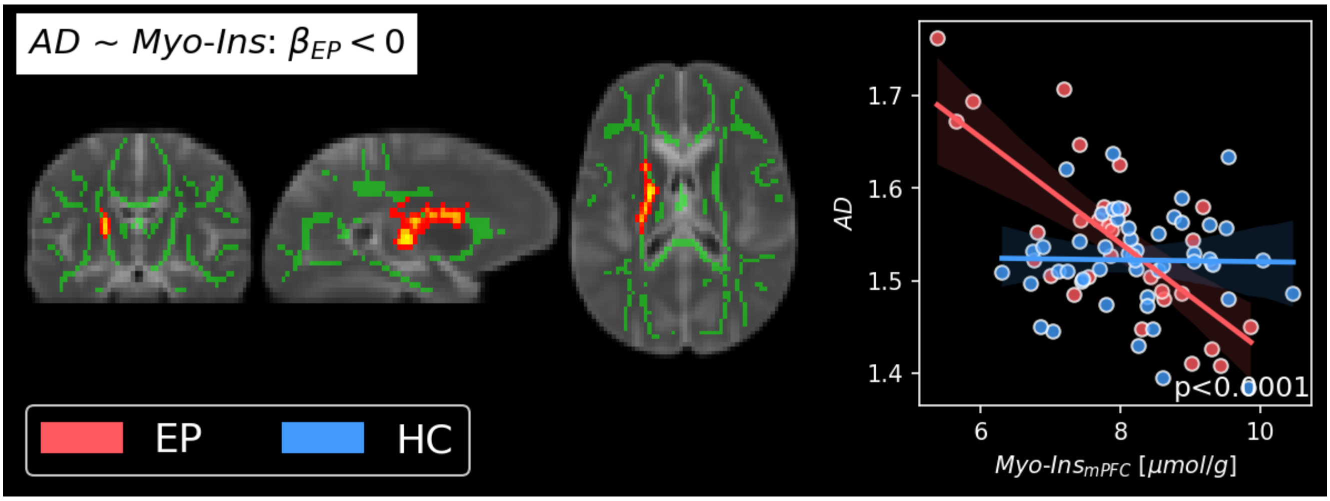


Figure S 1: Association clusters between myo-Ins of the mPFC and the microstructure metrics AD. Localized in the left internal capsule, AD is lower at higher concentrations of myo-inositol in the mPFC ($myo - Ins_{mPFC}$) in EP participants. $\beta_{EP} > 0$: contrast testing the slope of EP is significantly positive or negative. L: left hemisphere.

#### Post Hoc Analyses

##### NAA

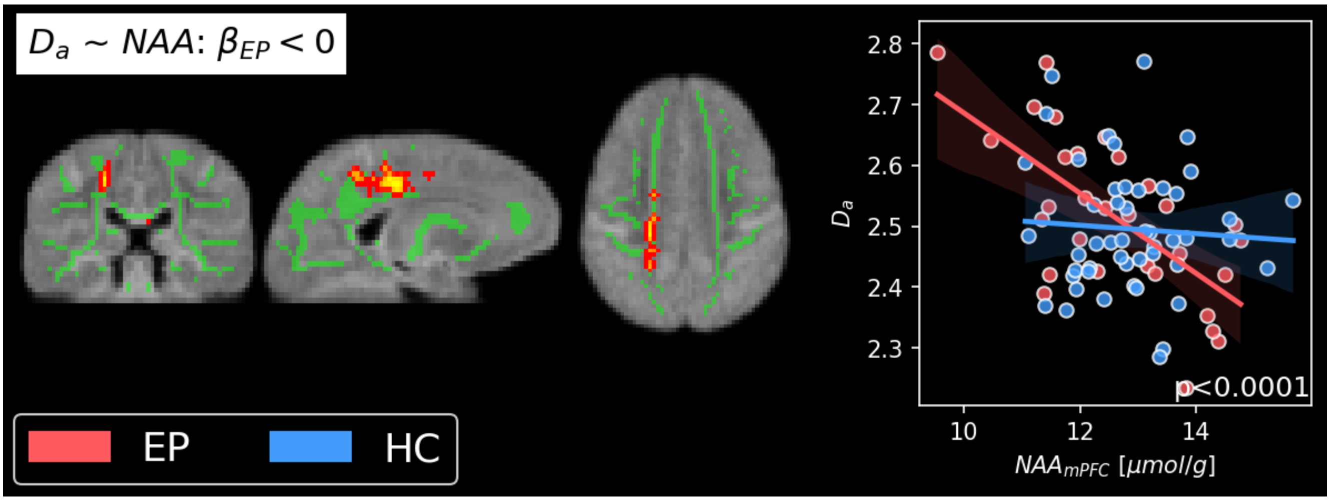


Figure S 2: TBSS clusters of associations between $D_a$ and NAA concentration in the mPFC. A cluster was found in parts of the left superior and posterior corona radiata, where Da was higher at lower concentrations of NAA ($NAA_{mPFC}$) in EP participants. In HC, no relationship with NAA and the WM metrics was found and the slope of EP did not significantly differ from HC. $\beta_{EP} < 0$: contrast testing the slope of EP is significantly negative.

##### Myo-inositol frequency mask

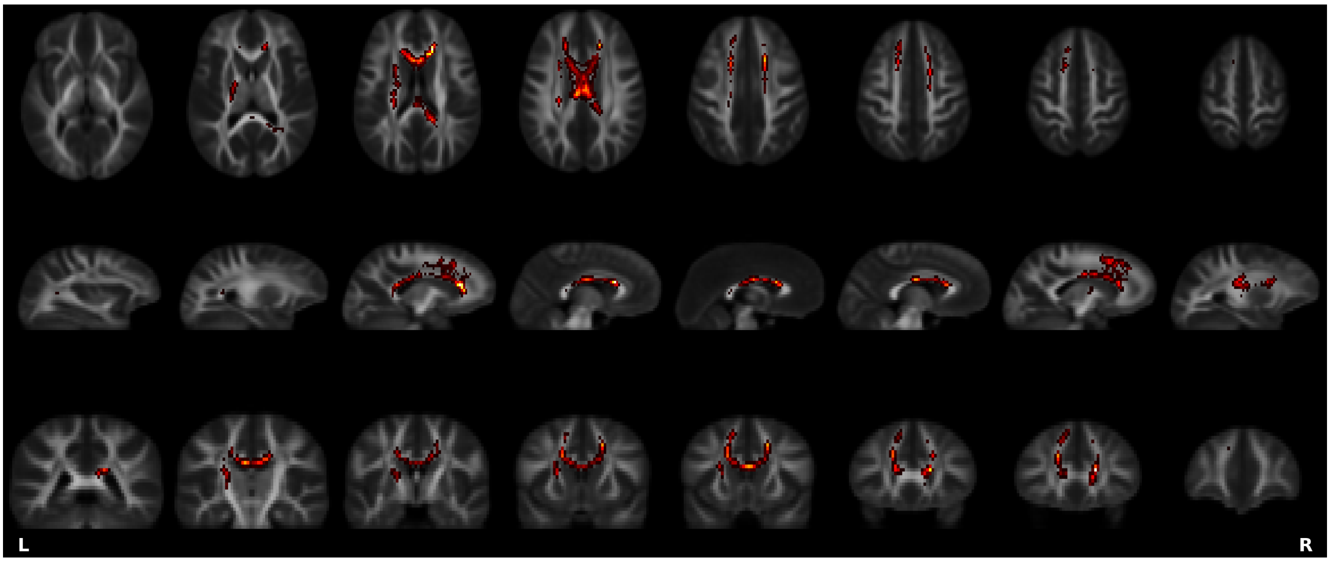


Figure S 3: WM mask used to define the cluster utilized in all post hoc analyses. The mask was generated by combining all the skeleton masks of significant WM clusters across dMRI modalities and contrasts of the myo-Ins associations. The mask comprises primarily forceps minor, region I, II and III of the corpus callosum (Hofer and Frahm (2006), i.e. genu and anterior part of the body), and anterior corona radiata. All these white matter regions contain projections and afferent fibers to the medial prefrontal cortex. Note, the mask was enlarged to aid the visualization with *tbss_fill*.

**Medication, duration of illness and Substance abuse**

In the literature a reduction in Glu and myo-In has been reported as a consequence of drug and/or cannabis exposure in non-psychiatric participants (Watts et al., 2020) and EP (Rigucci et al., 2018). In this cohort, no metabolites (all p$\geq$0.14) nor WM metric (all p$\geq$0.27) showed significant differences between EP participants exposed or not to cannabis. Similarly, alcohol and drugs consumption (CMRS total $\geq 1$) does not have an effect on WM metrics (p$\geq$0.72) or metabolites (p$\geq$0.14) in EP, except for GSH in which who consumes has higher GSH than who does not (p$\leq$0.03). None of the metabolites concentrations were associated to chlorpromazine equivalent dose (p$\geq$0.12) or duration of illness (p$\geq$0.35). Myo-Inositol WM clusters for $f$ and $D_{e,||}$ were not associated to chlorpromazine equivalent dose dose, p$\geq$0.16) or duration of illness (p$\geq$0.17).

### References

Y. Alemán-Gómez, T. Baumgartner, P. Klauser, M. Cleusix, R. Jenni, P. Hagmann, P. Conus, K. Q. Do, M. Bach Cuadra, P. S. Baumann, and P. Steullet. Multimodal Magnetic Resonance Imaging Depicts Widespread and Subregion Specific Anomalies in the Thalamus of Early-Psychosis and Chronic Schizophrenia Patients. *Schizophrenia Bulletin*, 49(1):196–207, Jan. 2023. ISSN 0586-7614, 1745-1701. doi: 10.1093/schbul/sbac113. URL `https://academic.oup.com/schizophreniabulletin/article/49/1/196/6692485`.

J. L. R. Andersson and S. N. Sotiropoulos. An integrated approach to correction for off-resonance effects and subject movement in diffusion MR imaging. *NeuroImage*, 125:1063–1078, Jan. 2016. ISSN 1095-9572. doi: 10.1016/j.neuroimage.2015.10.019.

S. Hofer and J. Frahm. Topography of the human corpus callosum revisited—Comprehensive fiber tractography using diffusion tensor magnetic resonance imaging. *NeuroImage*, 32(3): 989–994, Sept. 2006. ISSN 1053-8119. doi: 10.1016/j.neuroimage.2006.05.044. URL `https://www.sciencedirect.com/science/article/pii/S1053811906006501`.

E. Kellner, B. Dhital, V. G. Kiselev, and M. Reisert. Gibbs-ringing artifact removal based on local subvoxel-shifts. *Magnetic Resonance in Medicine*, 76(5):1574–1581, 2016. ISSN 1522-2594. doi: 10.1002/mrm.26054. URL `https://onlinelibrary.wiley.com/doi/abs/10.1002/mrm.26054`. _eprint: https://onlinelibrary.wiley.com/doi/pdf/10.1002/mrm.26054.

H.-H. Lee, D. S. Novikov, and E. Fieremans. Removal of partial Fourier-induced Gibbs (RPG) ringing artifacts in MRI. *Magnetic Resonance in Medicine*, 86(5):2733–2750, 2021. ISSN 1522-2594. doi: 10.1002/mrm.28830. URL `https://onlinelibrary.wiley.com/doi/abs/10.1002/mrm.28830`. _eprint: https://onlinelibrary.wiley.com/doi/pdf/10.1002/mrm.28830.

S. Rigucci, L. Xin, P. Klauser, P. S. Baumann, L. Alameda, M. Cleusix, R. Jenni, C. Ferrari, M. Pompili, R. Gruetter, K. Q. Do, and P. Conus. Cannabis use in early psychosis is associated with reduced glutamate levels in the prefrontal cortex. *Psychopharmacology*, 235(1):13–22, Jan. 2018. ISSN 1432-2072. doi: 10.1007/s00213-017-4745-z. URL `https://doi.org/10.1007/s00213-017-4745-z`.

C. M. W. Tax, M. Bastiani, J. Veraart, E. Garyfallidis, and M. Okan Irfanoglu. What's new and what's next in diffusion MRI preprocessing. *NeuroImage*, 249:118830, Apr. 2022. ISSN 1053-8119. doi: 10.1016/j.neuroimage.2021.118830. URL `https://www.sciencedirect.com/science/article/pii/S1053811921011010`.

J. Veraart, D. S. Novikov, D. Christiaens, B. Ades-Aron, J. Sijbers, and E. Fieremans. Denoising of diffusion MRI using random matrix theory. *NeuroImage*, 142:394–406, Nov. 2016. ISSN 1095-9572. doi: 10.1016/j.neuroimage.2016.08.016.

J. J. Watts, R. Garani, T. Da Silva, N. Lalang, S. Chavez, and R. Mizrahi. Evidence That Cannabis Exposure, Abuse, and Dependence Are Related to Glutamate Metabolism and Glial Function in the Anterior Cingulate Cortex: A 1H-Magnetic Resonance Spectroscopy Study. *Frontiers in Psychiatry*, 11:764, Aug. 2020. ISSN 1664-0640. doi: 10.3389/fpsyt.2020.00764. URL `https://www.ncbi.nlm.nih.gov/pmc/articles/PMC7468488/`.
